# Lowering catheter-associated urinary tract infections (CATION study) Statistical Analysis Plan

**DOI:** 10.64898/2026.06.24.26356490

**Authors:** Brett Mitchell, Philip Russo, Nicole White, Allen Cheng, Peta Tehan, David Brain, Katrina Browne, Georgia Matterson, Jennie King, Sally Havers

## Abstract

The CATION study is a parallel two-arm randomised controlled trial on the prevention of catheter-associated urinary tract infections (CAUTI) in hospitalised patients. The intervention is the use of a sterile wipe containing 0.1% chlorhexidine solution for meatal cleaning prior to urinary catheter insertion as part of usual care; the intervention will be compared with the use of a sterile wipe containing 0.9% normal saline as the control.

This document is the Statistical Analysis Plan for evaluating primary, secondary and tertiary effectiveness outcomes. The trial was preregistered on the Australian and New Zealand Clinical Trials registry (ACTRN12625000278437). A copy of the study protocol and a signed version of this Statistical Analysis Plan are available on request from the corresponding author (BM).

## STUDY SYNOPSIS

The CATION study is a parallel two-arm randomised controlled trial on the prevention of catheter-associated urinary tract infections (CAUTI) in hospitalised patients. The trial arms are a 0.9% normal saline (the control exposure) versus a 0.1% chlorhexidine solution (the intervention exposure) sterile wipe for meatal cleaning prior to urinary catheter insertion as part of usual care. Three hospitals are enrolled in the study, with randomisation occurring at participants’ time of study enrolment. The proposed study flowchart is in Figure 1.

**Figure 1:**
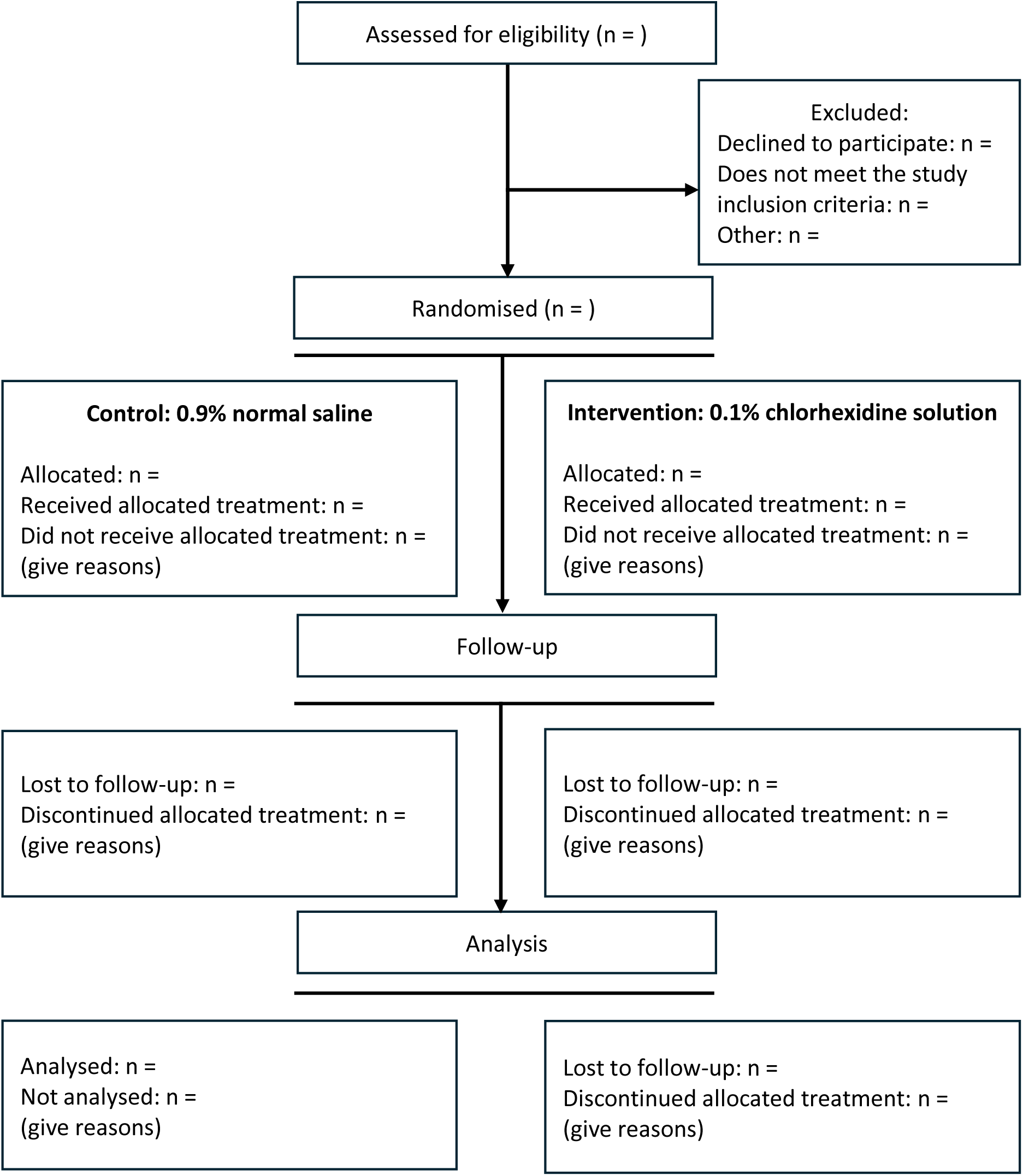
CONSORT study flowchart.

## STUDY OBJECTIVES

### PRIMARY OBJECTIVE

To compare confirmed CAUTI between hospitalised patients randomised to receive 0.1% chlorhexidine solution (intervention arm) versus 0.9% normal saline (control arm) for meatal cleaning prior to catheter insertion. This comparison will test if CAUTI incidence is equal (null hypothesis) or not equal (alternative hypothesis) between the intervention and control arms. The clinical hypothesis is that the use of 0.1% chlorhexidine solution will reduce CAUTI, compared with 0.9% normal saline.

### SECONDARY OBJECTIVES

To compare the intervention and control arms with respect to:

- Catheter-associated asymptomatic bacteriuria (CA-ASB)
- Time to first CAUTI from randomisation

The cost-effectiveness of the intervention compared with the control will also be evaluated. This objective will be outlined in a separate Health Economic Analysis Plan.

## PATIENT POPULATION

### INCLUSION CRITERIA

- Admitted as an inpatient at a trial hospital during the study timeframe, and
- Aged 18 years or over at study enrolment, and
- Receiving an indwelling urinary catheter as part of their usual medical care.

### EXCLUSION CRITERIA

- Aged less than 18 years at study enrolment and/or,
- Have an allergy, contraindication or other medical reason preventing the use of 0.1% chlorhexidine solution for cleaning the urethral meatal area and/or,
- Require in-and-out or suprapubic catheterisation and/or,
- Have symptoms and signs suggestive of, and/or are already receiving treatment for, urinary tract infection.

## OUTCOMES

### PRIMARY OUTCOME

Confirmed CAUTI in hospitalised patients from randomisation to follow-up. Follow-up is defined as 7 days post-catheter insertion, hospital discharge, or 48 hours post-catheter removal, whichever occurs first. The European Centres for Disease Control definition for CAUTI will be used to define confirmed cases:

1. The patient had an indwelling urinary catheter that had been in place for > 2 days on the date of event (day of device placement = Day 1) AND was either present for any portion of the calendar day on the date of event or removed the day before the date of event, and
2. The patient has at least one of the following signs of symptoms with no other recognised cause: fever (>38°C), urgency, frequency, dysuria, or suprapubic tenderness, and
3. The patient has a positive urine culture, that is, ≥ 10^5^ microorganisms per ml of urine with no more than two species of microorganisms or at least one of the following:

- positive dipstick for leukocyte esterase and/or nitrate;
- pyuria urine specimen with ≥10 white blood cells (WBC)/ml or ≥ 3 WBC/high-power field of unspun urine;
- organisms seen on Gram stain of unspun urine;
- at least two urine cultures with repeated isolation of the same uropathogen (Gram-negative bacteria or *S. saprophyticus*) with ≥10^2^ colonies/ml urine in non-voided specimens; − ≤10^5^ colonies/ml of a single uropathogen (Gram-negative bacteria or *S. saprophyticus*) in a patient being treated with effective antimicrobial agent for a urinary infection;
- physician diagnosis of a urinary tract infection;
- physician institutes appropriate therapy for a urinary infection.

A patient must meet all criteria (1 AND 2 AND 3) to meet the case definition.

The main analysis will define the primary outcome as the proportion of patients with confirmed CAUTI from randomisation to follow-up. The number of confirmed CAUTI per 100 catheter days will also be considered (see ‘Additional analyses’ section).

### SECONDARY OUTCOMES

Time to first CAUTI from randomisation to follow-up will be examined as a secondary outcome. This secondary outcome will be estimated as the cumulative incidence of CAUTI from randomisation to follow-up. Repeated events will not be included in the analysis.

The proportion of patients with CA-ASB from study enrolment to follow-up will be examined as a secondary outcome. A CA-ASB is defined as the presence of 10^5^ or more colony forming units of 1 or more bacterial species in a single catheter urine specimen, in a patient without symptoms that are compatible with a urinary tract infection.

Follow-up for secondary outcomes is defined as per the primary outcome definition (7 days post-catheter insertion, hospital discharge, or 48 hours post-catheter removal, whichever occurs first).

### TERTIARY OUTCOMES

Combined CAUTI and CA-ASB from randomisation to follow-up, whichever occurs first, will be examined as a secondary outcome. Tertiary outcomes will be analysed as time to first event (CA-ASB or CAUTI).

### SAFETY OUTCOMES

Adverse events will be monitored and documented through the trial period.

## INTERVENTION

The intervention being assessed is the use of a sterile wipe, containing 0.1% chlorhexidine solution, for meatal cleaning prior to urinary catheter insertion. The intervention will be compared with a control arm. The control arm is defined as the use of a sterile wipe, containing 0.9% normal saline, for meatal cleaning prior to urinary catheter insertion.

The allocated treatment from randomisation will be delivered by a dedicated hospital personnel, who has been trained and assessed as competent in catheter insertion by a study Chief Investigator. The hospital personnel will be Registered Nurses and will be available five days a week to insert catheters and follow up enrolled participants at each trial hospital. Known allergies will be checked prior to catheter insertion.

## RANDOMISATION AND BLINDING

Randomisation will be completed at the patient-level using the REDCap randomisation module. Eligible patients will be randomised to the intervention arm or the control arm at study enrolment. Randomisation will be stratified by hospital to balance treatment allocation by hospital. A block size of 6 will be used to balance allocation to control versus intervention arms within each hospital This is a double-blinded study, meaning that both the patient receiving the catheter and hospital personnel responsible for catheter insertion will be blinded to treatment allocation. Study data collectors will be blinded to treatment allocation to further reduce bias.

Allocation concealment will be achieved by labelling all products with a de-identifiable code, to ensure that neither the eligible patient nor hospital personnel will know whether the product being used is saline or chlorhexidine.

Only the principal investigator and one other chief investigator will have access to treatment allocations based on de-identifiable codes, needed to identify the intervention and control arms for analysis. The principal investigator and that chief investigator will not be involved in data collection or analysis. The mapping of de-identifiable codes to treatment allocations will be kept as a separate password-protected file. Only the principal investigator has access to this.

Once data collection is complete and prior to hand-over to the trial statistician, the assigned treatment arm for each participant will be labelled as “A” or “B” in the dataset. This labelling means that the trial statistician will be blinded when completing the planned analyses. Treatment allocation will be unblinded once analysis is complete and agreed to by all study investigators.

## SAMPLE SIZE

The target sample size was defined before study commencement, based on three enrolled hospitals. Calculations used local estimates for CAUTI incidence, estimated effectiveness informed by previous studies and available resources for participant recruitment.

CAUTI incidence in the control arm is estimated to be 6-7%. A previous multi-centre trial estimated a 74% reduction in the incidence of catheter-associated asymptomatic bacteriuria, and a 94% reduction in CAUTI (1). Conservative effect sizes of 70 – 90% were considered when estimating expected power under different control arm incidence estimates (Figure 2).

**Figure 2:**
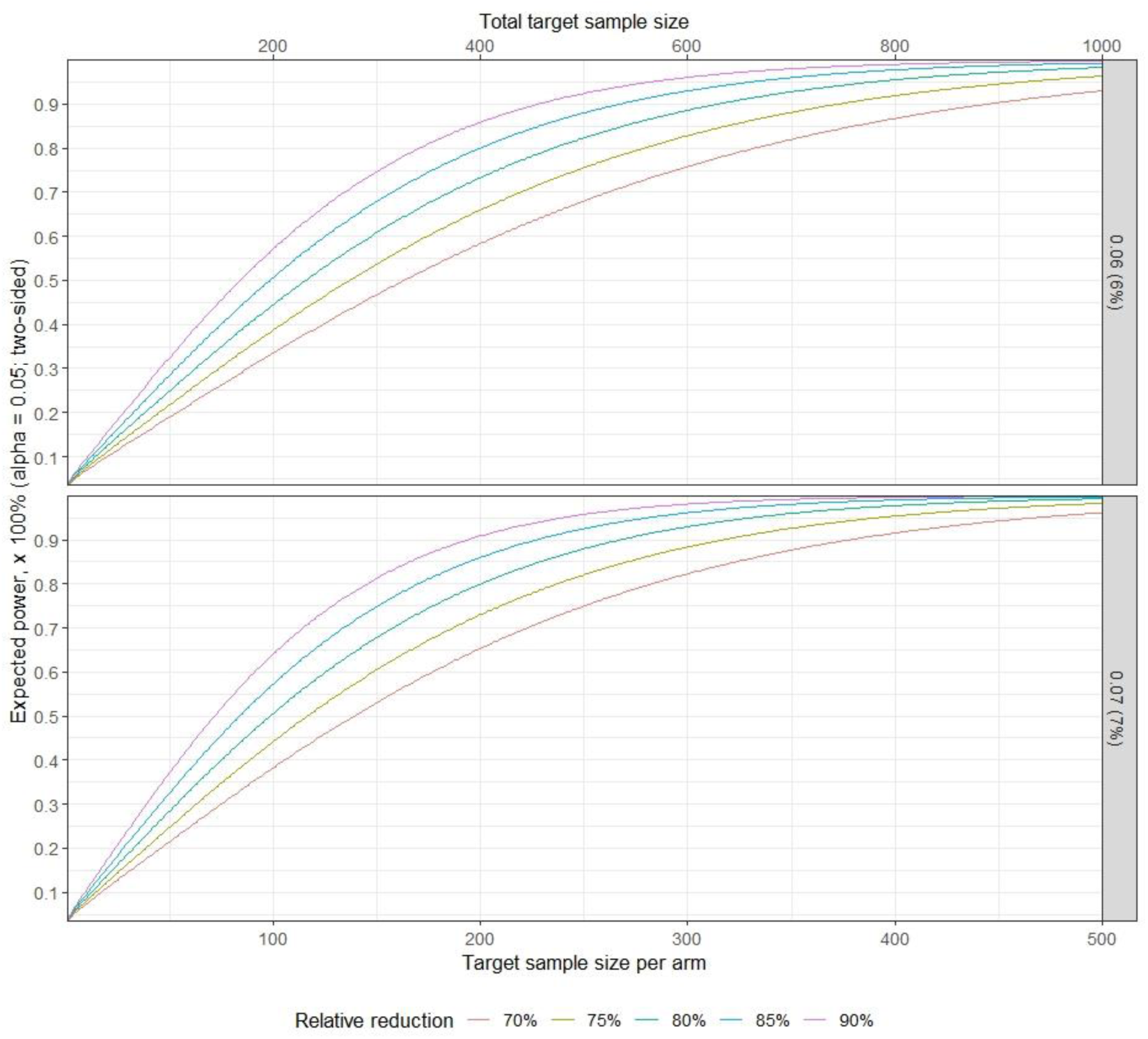
Expected power curves as a function of minimum sample size per trial arm (primary x-axis). The total target sample size is provided as a secondary x-axis at the top of the figure. The colours represent the estimated power for different relative reductions (%) in the primary outcome associated with the intervention arm compared with the control arm. Top panel: 6% incidence under the control arm; Bottom panel: 7% incidence under the control arm.

Expected power calculations were based on a two-sided alternative hypothesis with statistical significance defined at alpha = 0.05.

A 90% reduction in CAUTI with a sample size of 342 (171 each arm) would provide 80% power with a baseline incidence of 6%, assuming a 90% reduction. With the same power and baseline incidence, a 75% reduction would require a sample size of 558 (279 each arm).

Loss to follow-up and stratification by hospital were not accounted for in the calculation of expected power. Target sample size calculations are therefore conservative because they do not account for the percentage of variation in the primary outcome attributable to between-hospital differences (i.e., a variation-reduction factor is not applied); analyses will include stratification by hospital. Loss to follow-up is expected to be low, as eligible participants will be hospital inpatients who will be recruited and followed up while still in hospital.

## STATISTICAL ANALYSIS

### GENERAL PRINCIPLES

The scope of this Statistical Analysis Plan is the analysis of the primary outcome and secondary outcomes listed in this plan (see ‘Outcomes’ section). No changes have been made compared to the protocol (version 1.2) and information provided in the trial pre-registration on the Australia and New Zealand Clinical Trials Registry (ACTRN12625000278437). No interim analyses will be conducted.

The R statistical software (version 4.6.0 or higher) will be used to prepare data for analysis and to conduct primary and secondary outcome analyses. Analysis code and corresponding results will be collated into an R Markdown document and compiled as an HTML or Microsoft Word document for sharing with the study investigators.

The data will undergo additional cleaning by the trial statistician before starting data analysis. The purpose of this additional cleaning is to identify data entry errors that were not captured by rules encoded into the REDCap database used for data collection. Standard data cleaning processes will include identifying potential duplicate records, missing data checks and querying implausible values for date-time variables (e.g., a participant’s recorded follow-up date preceding their recorded date of randomisation) and incorrect values for patient-level variables based on the exclusion criteria (e.g., age less than 18 years). REDCap record identifiers associated with flagged records, if any, will be shared by the trial statistician with the research administrator and/or principal investigator for validation.

Records with missing data needed to define the follow-up period will be excluded. This exclusion and other exclusions based on data quality checks will be reported in the study CONSORT flowchart (Figure 1).

All analyses will be conducted on the intention-to-treat population only. Descriptive statistics for continuous variables will be expressed as mean and standard deviation, and quantile (Q1, Q2 [Median], Q3). Frequencies and percentages will be used to summarise categorical variables; denominators will be reported for variables with missing data.

Effect sizes associated with intervention exposure will be reported as odds ratios (OR) and incidence rate ratios (IRR) for binary outcomes and hazard ratios (HR) for time-to-event outcomes. Incidence rate ratios will be estimated from fitted model output using marginal standardisation. All effect sizes will be reported as point estimates with 95% confidence intervals.

Hypothesis testing will be conducted for the primary and secondary outcomes under the null hypothesis of no intervention effect. All hypothesis tests will assume a two-sided alternative hypothesis and a 5% statistical significance level. Hypothesis testing will not be applied to baseline characteristics.

The main analysis for primary and secondary outcomes will include stratification by hospital. Additional analyses will consider including patient-level variables collected at baseline and outcome assessment.

Results of all planned analyses, including figures and tables, will be presented as an Rmarkdown/Quarto report for investigator review and feedback on presentation.

### MULTIPLICITY ADJUSTMENT

There will be no multiplicity adjustment for primary and secondary outcomes, as secondary outcomes are only supportive of the primary outcome (CA-ASB) or are an alternative definition of the primary outcome (time to first CAUTI)

### BLIND REVIEW

A blind review of analysis results is planned, whereby the trial statistician and investigators will be blinded to the intervention and control arms via a random label (“A” or “B”). The purpose of the blind review is to obtain agreement from all investigators on the presentation of study findings.

Once full agreement is obtained, treatment allocation groups in the dataset will be updated to ‘Intervention’ and ‘Control’ and the analysis report rerun to produce the final results.

### DATA SETS TO BE ANALYSED

The intention-to-treat population will be analysed. The intention-to-treat population will include all randomised participants and will be analysed according to their allocated treatment group at randomisation.

A per-protocol analysis will not be conducted.

### SUBJECT DISPOSITION

The proposed CONSORT flowchart for participants is given by Figure 1. A second flowchart will report on the number of hospitals approached to participate in the study and the number of hospitals enrolled.

### PATIENT CHARACTERISTICS AND BASELINE COMPARISONS

Baseline characteristics will be summarised for the following participant groups:

- All eligible participants in the intention-to-treat population
- By hospital
- By treatment allocation (control, intervention)
- By primary outcome (no CAUTI, CAUTI) Baseline characteristics include:
- Age
- Sex
- Pre-existing comorbidities; Charlson comorbidity index
- Receiving antibiotics at time of catheter insertion
- Time from admission to catheter insertion (days)
- Type of catheter inserted

At outcome assessment, the following variables will be collected in addition to primary and secondary outcomes:

- Catheterisation duration (days)
- Reason for censor/end of follow up (death, discharge, 7 days or catheter removal)

### COMPLIANCE TO STUDY INTERVENTION(S)

Intervention compliance is expected to be complete because treatment is administered at the time of catheter insertion. Any deviations from allocated treatment will be reported.

## STATISTICAL ANALYSIS

### PRIMARY OUTCOME MAIN ANALYSIS

The primary outcome is confirmed CAUTI during the defined follow-up period (within 7 days of catheter insertion, hospital discharge, or 48 hours post-catheter removal, whichever occurs first). The dependent variable will be analysed as binary, equal to 1 for confirmed CAUTI and 0 otherwise. The main analysis estimand is the conditional effect of the intervention, which will be estimated by the odds ratio (OR)

A logistic regression model will estimate the odds of CAUTI associated with the intervention arm compared with the control arm. The treatment arm will be defined as a binary fixed effect, equal to 0 for the control arm and 1 for the intervention arm. Stratification by hospital will be accounted for in the regression model by including binary dummy variables for two of the three hospitals; the remaining hospital will be set to zero for identifiability.

Nonparametric bootstrapping will be used to estimate the difference in proportions and the relative risk between intervention and control arms, using the fitted logistic regression model. The results will represent marginal estimands for the intervention effect.

### SECONDARY OUTCOME ANALYSIS

#### CA-ASBs

The analysis will follow the same steps as per the primary outcome.

The presence of CA-ASB during participant follow-up will be defined as a binary dependent variable (0 = CA-ASB not present, 1 = CA-ASB present). This secondary outcome will be analysed using logistic regression with a binary fixed effect for the intervention (0 = Control, 1 = Intervention) and fixed effects to account for stratification by hospital.

#### Time to first CAUTI

Time to first CAUTI will be described using Kaplan-Meier survival curves. A log-rank (Mantell-Cox) test will be used to compare survival distributions for control and intervention arms. A Cox proportional hazard model will be developed to estimate the hazard of CAUTI (yes or no), comparing those in the control and intervention arms. The proposed null hypothesis is that use of chlorhexidine 0.1% will not increase the hazard of a CAUTI compared to saline 0.9%. Hazard ratios will be adjusted for age, sex, and other clinically relevant patient characteristics assessed at baseline. The proportional hazards assumption will be assessed by Schoenfeld residuals. If there is strong evidence of non-proportional hazards, a time-dependent intervention effect and/or non-parametric models will be considered.

### TERTIARY/EXPLORATORY ANALYSIS

Combined CAUTI and CA-ASB will be analysed as a tertiary outcome, as detailed in the methods for the main analysis. In the event that a patient records both CA-ASB and CAUTI within the follow-up period, this will count as a single event to avoid double-counting.

This main analysis assumes that catheter duration is the same between the control and intervention arms. If the trial data indicate that catheter duration differs by trial arm, the primary outcome will also be analysed using a Poisson regression model with catheter days divided by 100 as a model offset. The corresponding incidence rate ratio will estimate CAUTI per 100 catheter days.

To explore the potential influence of disease severity at randomisation on outcomes, stratification by the Charlson Comorbidity Index will be considered as an exploratory analysis.

### SUBGROUP ANALYSES

A per-hospital analysis will be conducted for both primary and secondary outcomes, by fitting the defined models to each hospital separately.

### TREATMENT OF MISSING DATA

Given the inpatient setting, missing outcome data are expected to be minimal. The extent of missing outcome data will be reported. In the event of missing data, multiple imputation will be used to estimate missing covariates included in the adjusted analysis of primary and secondary outcomes.

Candidate covariates will only be imputed if data are missing for less than 10% of the intention-to-treat population. Multiple Imputation using Chained Equations will be used; the expected number of iterations to achieve convergence is 10 – 20 iterations. Participants with missing outcomes will be excluded from analysis.

### ADDITIONAL ANALYSES

For binary outcomes, a binomial model with a log link will be considered as a sensitivity analysis, subject to model convergence. The log link will estimate the relative risk of each outcome that is associated with the intervention arm versus the control arm.

### SAFETY OUTCOME ANALYSIS ADVERSE EVENTS

Any adverse events reported during the study will be reported descriptively. No formal analysis or comparison between study arms will be undertaken.

## Data Availability

All data produced in the present work are contained in the submission

## LIST OF ABBREVIATIONS

CA-ASB: Catheter-associated asymptomatic bacteriuria
CAUTI: Catheter-associated urinary tract infection
HR: Hazard ratio
OR: Odds ratio
RR: Relative risk
WBC: White blood cells

